# Markers of environmental enteric dysfunction are associated with changes in pharmacokinetics of praziquantel in preschool age children with *Schistosoma mansoni* infection in Albertine region of Uganda

**DOI:** 10.64898/2026.09.02.26362031

**Authors:** Andrew Edielu, Bonniface Obura, Patrice A. Mawa, Martin J. Holland, Emily L. Webb, Alison M. Elliott, Gloria Kakoba Ayebazibwe, Hannah Wei Wu, Nick Mancini, Fabian C. Fischer, Susannah Colt, Meagan A. Barry, William Hope, Catriona Waitt, Jennifer F. Friedman, Amaya L. Bustinduy

## Abstract

**Introduction:** Praziquantel (PZQ) is the only widely available chemotherapy that is effective against all species of schistosomes. Environmental enteric dysfunction (EED) is an acquired intestinal disorder of altered gut function whose effect on drug pharmacokinetics has not been directly explored.

**Methods:** Preschool-age children infected with *S. mansoni* were randomized to receive 40mg/kg or 80mg/kg of crushed PZQ tablets. Plasma PZQ concentrations were quantified using ultra-high performance liquid chromatography mass spectrometry. Maximum concentration (C_max_), time to C_max_ (T_max_) and area under the curve (AUC) of PZQ were calculated. Biomarkers of intestinal inflammation (stool calprotectin), epithelial damage (plasma Intestinal Fatty Acid Binding Protein (IFABP)), permeability (urine lactulose:mannitol (LM) ratio and alpha-1 antitrypsin (AAT)), microbial translocation (plasma Endotoxin core antibodies (EndoCAb), systemic inflammation (plasma C-reactive protein (CRP)), and presence of faecal occult blood (FOB) were measured. Using linear regression, we assessed association of AUC, C_max_, T_max_ and R- to S-PZQ exposure with each biomarker, adjusting for dose, age, and sex.

**Results:** Of the 184 participants included in the final analysis, 91 received 40mg/kg and 93 received 80mg/kg of PZQ. The T_max_ was associated with LM ratio (β=0.06, 95% CI 0.02 – 0.11, p=0.003) and calprotectin (β=0.001, 0.0002 – 0.002, p=0.013). CRP was associated with AUC (β=0.13, 95% CI 0.06 – 0.21, p=0.001) and C_max_ (β=0.12, 95% CI 0.05 – 0.21, p=0.002). Calprotectin (β=0.12, 95% CI 0.04 - 0.20, p=0.003) and AAT (β=0.09, 95% CI 0.02 – 0.15, p=0.008) were associated with a higher R-PZQ/S-PZQ AUC ratio, while CRP was not (p=0.38).

**Conclusion:** Elevated intestinal inflammatory markers were associated with increased T_max_, indicating reduced rate of absorption and relative increase in exposure to the active R-enantiomer. Systemic inflammation was associated with higher C_max_ and AUC, implying increased exposure to PZQ. This is the first report linking alterations in praziquantel pharmacokinetics to EED markers and systemic inflammation.

**Author summary:** Schistosomiasis, also known as bilharzia, affects approximately 240 million people in 79 countries with children in Africa bearing almost 50% of the burden. Praziquantel (PZQ), an oral drug, is the only widely available treatment for the disease. However, some studies have reported differences in pharmacokinetics and efficacy of PZQ across geographical locations suggesting environmental factors to affect drug response. We hypothesized that environmental enteric dysfunction, a disorder of altered intestinal function, may affect absorption and metabolism of PZQ because of the damage it inflicts on the intestinal mucosa, and subsequent systemic inflammation due to leakage of intestinal contents, a process referred to as “microbial translocation”. Our findings indicate that markers of intestinal inflammation were associated with reduced rate of absorption while systemic inflammation was associated with increased plasma concentrations and overall exposure to PZQ. The reduced rate of absorption may result from destruction of the intestinal absorptive surface while increased plasma concentration and exposure may be due to reduced activity of PZQ-metabolizing enzymes in the liver. These observations could inform approaches for optimizing the effectiveness of PZQ by accounting for inflammation. This has implications for the use of PZQ as an effective control strategy and subsequent elimination of schistosomiasis by 2030, in line with World Health Organization targets.

## Introduction

Schistosomiasis affects approximately 240 million people in 79 countries and is caused by a trematode of the genus *Schistosoma (S.),* with human disease mainly attributed to *S. mansoni, S. japonicum* and *S. haematobium*. Most of the burden of the disease is in Africa, with high prevalence in preschool age children (PSAC) [1]. Mass drug administration (MDA) of praziquantel (PZQ), the only available treatment for schistosomiasis, remains the most commonly used control strategy. However, PSAC have until recently been excluded from such programs because PZQ is not licenced in this age group [2].

Praziquantel is a racemate composed of a biologically active enantiomer (R-PZQ) and an inactive distomer (S-PZQ). Following oral administration, PZQ is rapidly absorbed, reaching maximum plasma concentration in 2.0 - 2.6 hours. It is approximately 80% plasma protein-bound and undergoes extensive first-pass metabolism by cytochrome P450 (CYP) enzymes, yielding trans-4-hydroxypraziquantel as the principal metabolite. Metabolites are excreted renally, with 80% of the dose recovered in urine within 24 hours [3]. Most of the absorbed PZQ is metabolized by CYP enzymes CYP1A2, CYP3A4, CYP2D6, CYP3A5, and CYP2C19 [4], and voltage-gated calcium channels are the presumed targets for its schistosomicidal effect. Exposure to PZQ triggers uptake of calcium ions and vacuolation of the adult schistosome worms [5], followed by contraction and paralysis and, ultimately, immune-mediated clearance by the host [6].

Despite the lack of evidence of resistance to PZQ [7], variation in schistosomiasis cure rate has been reported across populations and geographical settings [8, 9]. High variability of efficacy of PZQ against *S. mansoni* has been observed in East Africa at 97.9% in Rwanda, 90.4% in Ethiopia, 75.4% in Tanzania, 71.2% in Kenya, and 68.4% in Uganda [10]. This variation has been attributed to factors such as concomitant food intake [11], genetic polymorphisms affecting CYP activity [12] as well as differences in infection burden [10]. Crushing PZQ tablets has also been shown to increase the rate of absorption in PSAC [13]. In comparison to older populations, cure rate is lower in PSAC using the standard 40mg/kg dose, which has been linked to lower systemic PZQ exposure [14]. Reduced PZQ efficacy has been demonstrated in malnourished mice [15], an effect that may be mediated, in part, by impaired absorption since intestinal inflammation and epithelial damage can impair oral drug absorption. This suggests that gut dysfunction may contribute to variable PZQ pharmacokinetics (PK) [3, 16]. Similarly, decreased efficacy of oral vaccines has been observed in the presence of intestinal inflammation albeit with mixed findings [17, 18].

Environmental enteric dysfunction (EED) is an acquired subclinical condition commonly attributed to chronic exposure to enteric pathogens because of poor water, sanitation and hygiene. It is characterized by intestinal inflammation and destruction of the epithelium which is associated with malabsorption due to decreased villous absorptive capacity. Disruption of the intestinal epithelium culminates in microbial translocation into the normally sterile bloodstream, with subsequent systemic immune activation. The malabsorption and continuous inflammation ultimately lead to undernutrition, which is considered a key outcome of EED [19].

EED may affect the PK of PZQ through several mechanisms. First, EED-associated malabsorption may reduce drug absorption [16]. Second, inflammation supresses the activity of hepatic CYP enzymes thereby increasing drug exposure [20, 21]. EED also has the potential to reduce immune function directly, and indirectly by inducing malnutrition. This leads to impaired schistosome clearance and lower efficacy [15, 18, 22].

Definitive diagnosis of EED requires intestinal biopsy which is impractical in field settings, particularly in young children [23]. Biomarkers are therefore used as proxies for the key pathologic features associated with EED including a) intestinal inflammation b) intestinal permeability c) enterocyte damage d) microbial translocation and systemic inflammation [24]. Considering that EED and schistosomiasis are endemic in similar settings, it is possible that EED contributes to the observed variation in PZQ PK and therapeutic response. In this study, we investigate the association of EED biomarkers with various PK outcomes of PZQ and offer insights into environmental factors that may affect the PK of PZQ, which may have implications for therapeutics for other conditions in the setting of comorbidity with EED and other inflammatory disorders.

## Methods

### Study population and setting

This study utilized data and stored baseline samples from the ‘*Praziquantel for children under age four years: A Phase II PK/PD driven dose finding trial (PIP trial)*’ [25, 26] which recruited PSAC aged 12-47 months infected with *S. mansoni* from fishing villages on the shores of Lake Albert in Western Uganda from February to December 2021. Participants with grade 3 or higher abnormality of renal or liver function, severe wasting, exposure to immunomodulatory drugs or significant illness from clinical assessment were excluded. Data from 184 PIP trial participants who participated in the pharmacokinetics sub-study were used, and PK analysis done only for the drug administered on the day of enrolment.

***Diagnosis of S. mansoni* infection**: Confirmation of schistosomiasis infection was done through sequential testing with urine circulating cathodic antigen (Schisto POC-CCA^®^ Rapid Medical Diagnostics, Pretoria, South Africa), followed by stool Kato-Katz technique and finally upconverting phosphor-lateral flow circulating anodic antigen (UCP-LF CAA) assay (Leiden University Medical Centre, Netherlands) as detailed previously [25]. Only participants with *S. mansoni* infection indicated by all the three tests were enrolled.

### Administration of Praziquantel

In the PIP trial [26], participants were randomized to receive 80mg/kg split into two doses of 40mg/kg three hours apart, or a single dose of 40mg/kg of praziquantel. Each tablet of PZQ of 600mg is scored three times, yielding four equal pieces. At each timepoint, a dose of 40mg/kg was used and the final amount administered was rounded to the nearest 150mg (one quarter of a tablet) as per recommended dosing for PSAC [27]. The pieces of tablet were crushed and the powder mixed with mango juice. The cup was handed to the mother or the child for the older participants. The dose was repeated if the child vomited within 30 minutes of administration. PZQ was administered just after breakfast or lunch.

### Sample and data collection and processing

Blood samples were collected in lithium heparin vacutainers at four time points, namely 1.5 hours, 3 hours, 4 hours and 6-8 hours after product administration. The samples were centrifuged for 15 minutes at 2500 RPM and the plasma pipetted into 2ml pre-labelled cryotubes and immediately stored in liquid nitrogen. They were then transferred to MRC/UVRI & LSHTM Uganda Research Unit, Entebbe, Uganda for longer-term storage at −80°C awaiting transportation to University of Liverpool, United Kingdom where the pharmacokinetic analysis was done or to the Center for International Health Research (CIHR) and Texas Children’s Hospital in the United States where quantification of biomarkers and lactulose:mannitol were done respectively.

### Chemical Analytics

Praziquantel plasma concentrations were quantified using a ultra-high pressure liquid chromatography system (Waters Acquity) coupled to a triple-quadrupole Waters TQ-XS mass spectrometer (LC-MS/MS) assay. The internal standards, d11 (R) and d11 (S) PZQ (Toronto Research Chemicals) were prepared in acetonitrile + 0.1% formic acid (Fisher Scientific UK) and 150 µL was added to a 96-well protein precipitation plate (Phenomenex, Cheshire, UK). Calibration and quality control samples were prepared from stock solutions of R-PZQ and S-PZQ (1 mg/mL; Toronto Research Chemicals) by serial dilutions in water and methanol (1:1 v/v) to obtain concentrations in the range 0.01 – 10 mg/L. 25 µL each of samples, blanks, quality control and calibration samples were mixed with the internal standard on an orbital shaker for 5-min at 600 rpm. Analyte was extracted through the protein precipitation plate into a collection plate using a positive pressure manifold. Water and acetonitrile containing 0.1% formic acid (200 µL) was added to each well. Collection plates were sealed, and mixing done on an orbital shaker prior to analysis by LC-MS/MS. Chromatographic separation was achieved by injection of analytes (3 µL) onto a Waters Trefoil CEL2 Column (2.5 μm, 2.1 mm X 50 mm) and separated over a 5 min gradient using a mixture of solvents A (LC-MS grade water + 0.1% formic acid) and B (LC-MS grade acetonitrile + 0.1% formic acid). Separations were accomplished by applying a linear gradient of 95% to 5% solvent A over 3.5 min at a flow rate of 0.4 mL/min followed by an equilibration step (1.5 min at 95% solvent A). The mass spectrometer was operated in positive ion mode and multiple reaction monitoring (MRM) method used for detection and quantification of R- and S-PZQ. The lower limit of quantification (LOQ) was 0.01 mg/L.

### Pharmacokinetic analysis

Noncompartmental analysis was used to determine R- and S-PZQ pharmacokinetic parameters. The maximum plasma concentration (C_max_; mg/L), time to reach C_max_ (T_max_; h), and area under the concentration-time curve (AUC) from 0 hours to the last observed measurable concentration (AUC_0-t_; mg*h/L) were estimated. The apparent elimination rate constant (k_e_) was estimated from the ln-linear regression of post-peak concentrations. The R/S exposure ratio (R-PZQ/S-PZQ) was calculated per participant for C_max_ and AUC. For the 40 mg/kg single-dose group, the absorption rate constant (k_a_) was estimated for each participant by numerically solving the one-compartment oral-absorption expression for T_max_ with Brent’s root-finding algorithm, and participants were classified as showing conventional (k_a_ > k_e_) or flip-flop (k_a_ < k_e_) absorption kinetics. The k_a_ estimates were considered reliable only for participants whose observed peak was predicted to occur after the first sampling time (T_max_ > 1.5 h, *n* = 86). The 80 mg/kg group was excluded from k_a_ estimation because the second dose at three hours superimposes on the first-dose profile. Full details of the enantiomer-specific NCA, the k_e_ and k_a_ derivations, and the equations used are provided in *Section S1* of the supplementary information.

### Assessment of EED biomarkers

Biomarkers representing various EED pathophysiological pathways [19] were assessed. Intestinal inflammation was assessed by quantifying calprotectin in fresh stool samples using Buhlmann Quantum Blue^®^ Lateral Flow Assay (Alpha Laboratories, Hampshire, United Kingdom). Urine lactulose:mannitol (LM) ratio was used as a marker of intestinal permeability. The LM test was conducted by administering a solution containing 250 mg/ml lactulose and 50 mg/ml mannitol in sterile water (constituted at Hospice Africa Uganda, Kampala, Uganda) at a dose of 2ml/kg up to a maximum of 20ml and all the urine voided in the two hours after ingestion of the solution was collected in a clean container with chlorhexidine to avoid bacterial overgrowth. The total urine volume was measured and two aliquots of 2ml each were collected and stored. Participants fasted for two hours prior to administration of the sugar solution and during urine collection, except for breastfeeding infants [24]. Quantification of lactulose and mannitol was done by high-performance liquid chromatography at Texas Children’s Hospital in Houston, Texas, USA [28]. Using the respective manufacturers’ manuals, ELISA was used for assessment of stool Alpha-1 antitrypsin (AAT) (BioVendor R&D, catalogue #IC6200) as a marker of intestinal permeability, plasma Intestinal Fatty Acid Binding Protein (I-FABP) (Hycult Biotech, catalogue #HK406-01) as a marker of enterocyte damage and plasma Endotoxin core antibody (EndoCAb) (Hycult Biotech, catalogue # HK504-IgG) as a marker of microbial translocation. A multiplex bead-based assay developed at CIHR, USA [29] was used to quantify plasma CRP as a marker of systemic inflammation. Presence of faecal occult blood (FOB), a marker of gastrointestinal bleeding was also qualitatively measured using a point-of-care rapid test cassette (Biopanda Reagents, United Kingdom).

### Statistical Analysis

All statistical analyses were performed in STATA v18.0 (StataCorp, Texas, USA). We tested the hypothesis that higher concentrations of EED biomarkers impact the PK of PZQ because of its influence on the absorptive surface. PK outcome variables were C_max_, T_max,_ AUC, k_a_, and incidence of flip-flip kinetics (k_a_ < k_e_). The independent variables considered were LM ratio, I-FABP, AAT, EndoCAb and Calprotectin which are EED biomarkers, as well as CRP, a systemic inflammatory marker.

Descriptive statistics were summarized for the variables under consideration, including demographics (age and sex). Kruskal-Wallis non-parametric test was done to compare the descriptive statistics of each continuous variable after grouping the observations by dose i.e. 40mg/kg versus 80mg/kg, while chi-squared test was used for categorical variables. Linear regression models were used for log transformed AUC and C_max_. The T_max_ was used in the model without transformation because measurements were done at specific time intervals for all participants, yielding blocks/clusters of measurements, rather than the typical continuous variables, as was the case with the other two outcome variables. The resulting distribution of T_max_ was closer to normal compared to the log-transformed variable.

Each PK outcome, namely AUC, C_max_ and T_max_, was regressed on each biomarker in a separate multivariable linear model adjusted for PZQ dose, with age and sex included as covariates (Section on supplementary information). Because both the outcome and the biomarker were log-transformed, each regression coefficient (β) is interpreted as the approximate percentage change in the PK outcome per 1% change in the biomarker. The potential role of the PZQ dose as an effect modifier was assessed for each model. A p-value of ≤0.05 was used to denote statistical significance. The same regression framework was additionally applied to the enantiomer-specific exposure parameters and to the R/S ratio in order to test whether biomarkers were associated with stereoselective disposition.

### Ethics statement

The PIP trial received ethics approval from the Uganda Virus Research Institute (No. GC/127/19/07/708), National Drug Authority (No. CTC 0133/2020), the Uganda National Council for Science and Technology (No. HS 2650), Rhode Island Hospital (No. 401020), and the London School of Hygiene & Tropical Medicine (No. 14851). Participants who were diagnosed with schistosomiasis but excluded due to other eligibility criteria received praziquantel.

## Results

### Baseline characteristics

A total of 190 PSAC who were recruited to the PK arm of the PIP trial, of which six were excluded from analysis because of incomplete data on the PK parameters. Of the 184 participants included in the final analysis, 91 (49.4%) received 40mg/kg of PZQ and a placebo three hours later while the rest received a total of 80mg/kg divided into equal 40 mg/kg doses three hours apart. Participants who received 40mg/kg did not differ from those who received 80mg/kg with respect to demographic characteristics or parasitological parameters as detailed in ***Table 1***.

**Table 1:**
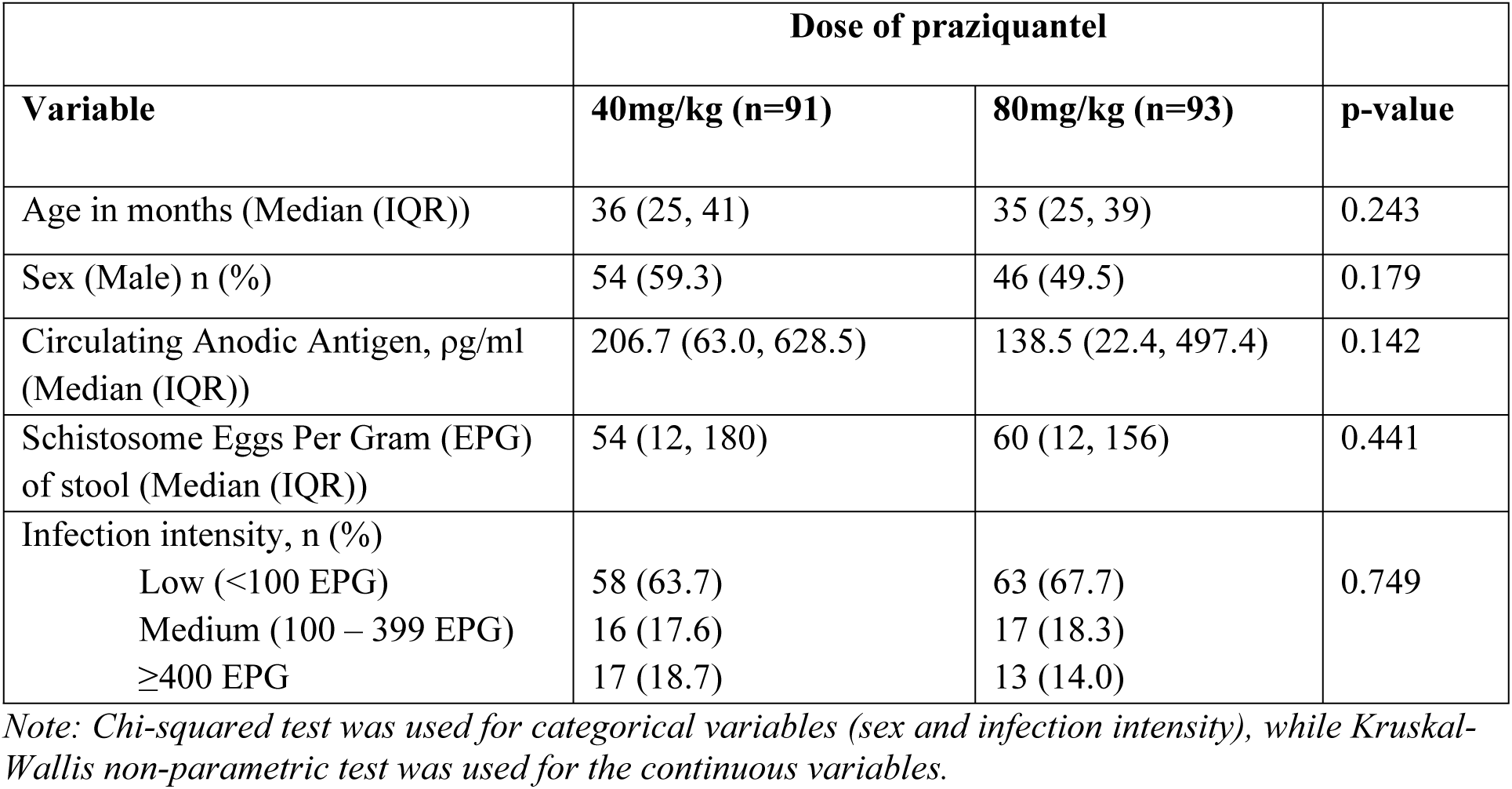
Baseline characteristics of PSAC that underwent praziquantel pharmacokinetic analysis after treatment for *S. mansoni* in Albertine Region of Western Uganda.

### Association of PK parameters with biomarkers

As expected, area under the curve (AUC) and C_max_ were significantly higher in the 80mg/kg group which is explained by a higher overall dose (p < 0.001). The median T_max_ was also higher in the 80 mg/kg group, also expected due to the three-hour interval between the two 40mg/kg doses given in this group ***(Table 2)*.**

**Table 2:** Praziquantel pharmacokinetic parameters in PSAC treated for *S. mansoni* in Albertine Region of Western Uganda.

|  | 40mg/kg (n=91)<br>Median (IQR) | 80mg/kg (n=93)<br>Median (IQR) | p-value |
| --- | --- | --- | --- |
| Maximum concentration ($C_{\max}$), mg/L | 0.11 (0.04, 0.16) | 0.33 (0.15, 0.65) | <0.001 |
| Time to maximum concentration ($T_{\max}$), hours | 2.8 (1.82, 4.00) | 4.00 (3.92, 6.08) | <0.001 |
| Area Under the Curve (AUC), mg*h/L<br>ng.h/mL | 0.32 (0.13, 0.46) | 0.71 (0.39, 1.38) | <0.001 |

Note: Kruskal-Wallis non-parametric test was used to compare the medians

The AUC was associated with elevated CRP levels (p-value = 0.001) (**Figure 1**). No other biomarker showed a significant association with AUC. The same trend was observed with C_max_ which was associated with significantly higher CRP (p-value = 0.002) but not associated with other biomarkers as shown in **Figure 2**.

**Figure 1:**
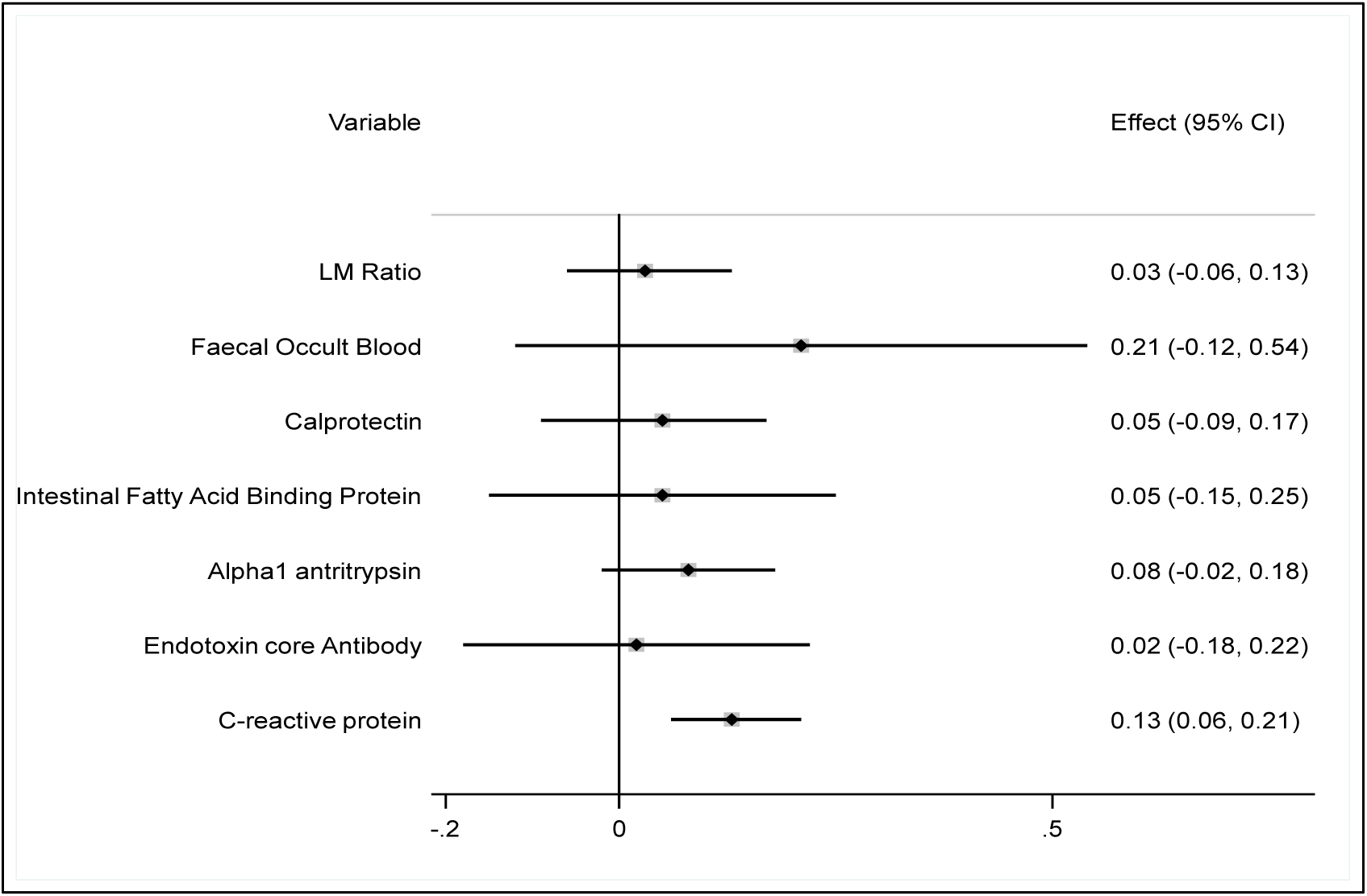
Forest plot showing association of log-transformed Area under the Curve (AUC) of praziquantel with EED biomarkers after adjusting for age, sex and dose of PZQ. For each variable in the column on the left, the β regression coefficients are represented by the horizontal lines with the box being the effect size and the line on either side the 95% confidence intervals (CI). This is further presented as numerical values in the column on the right. Variables whose 95% CI do not cross the zero line on the x-axis are statistically significant.

**Figure 2:**
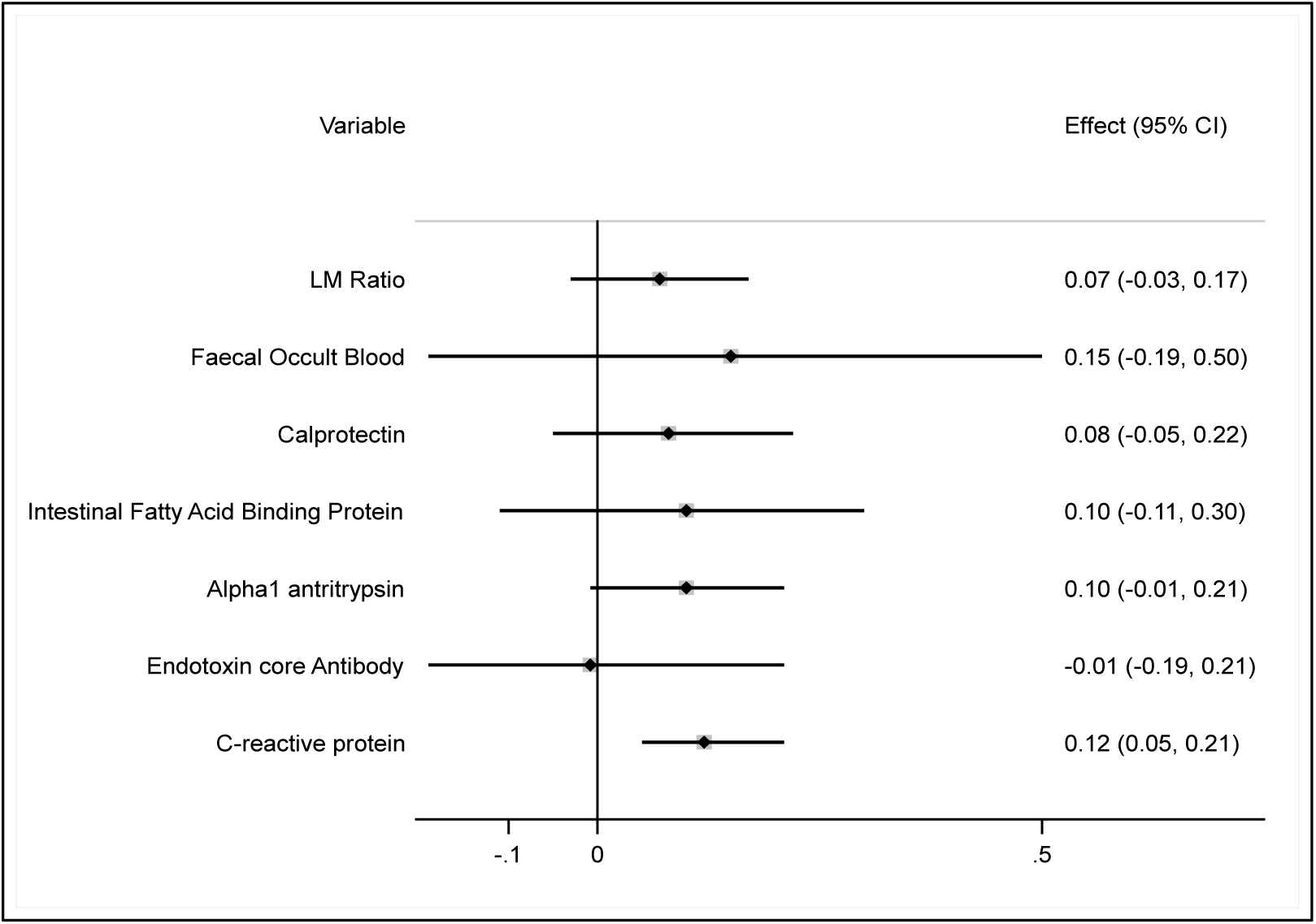
Forest plot showing association of log-transformed plasma maximum concentration (C_max_) of praziquantel with EED biomarkers after adjusting for age, sex and dose of PZQ. For each variable in the column on the left, the β regression coefficients are represented by the horizontal lines with the box being the effect size and the line on either side the 95% confidence intervals (CI). This is further presented as numerical values in the column on the right. Variables whose 95% CI do not cross the zero line on the x-axis are statistically significant.

The T_max_ was associated with LM ratio (p-value = 0.003) and calprotectin (p-value=0.013). The positive values of the β coefficients implies that elevated levels of the two biomarkers are associated with longer time to maximum concertation **(*Figure 3*).**

**Figure 3:**
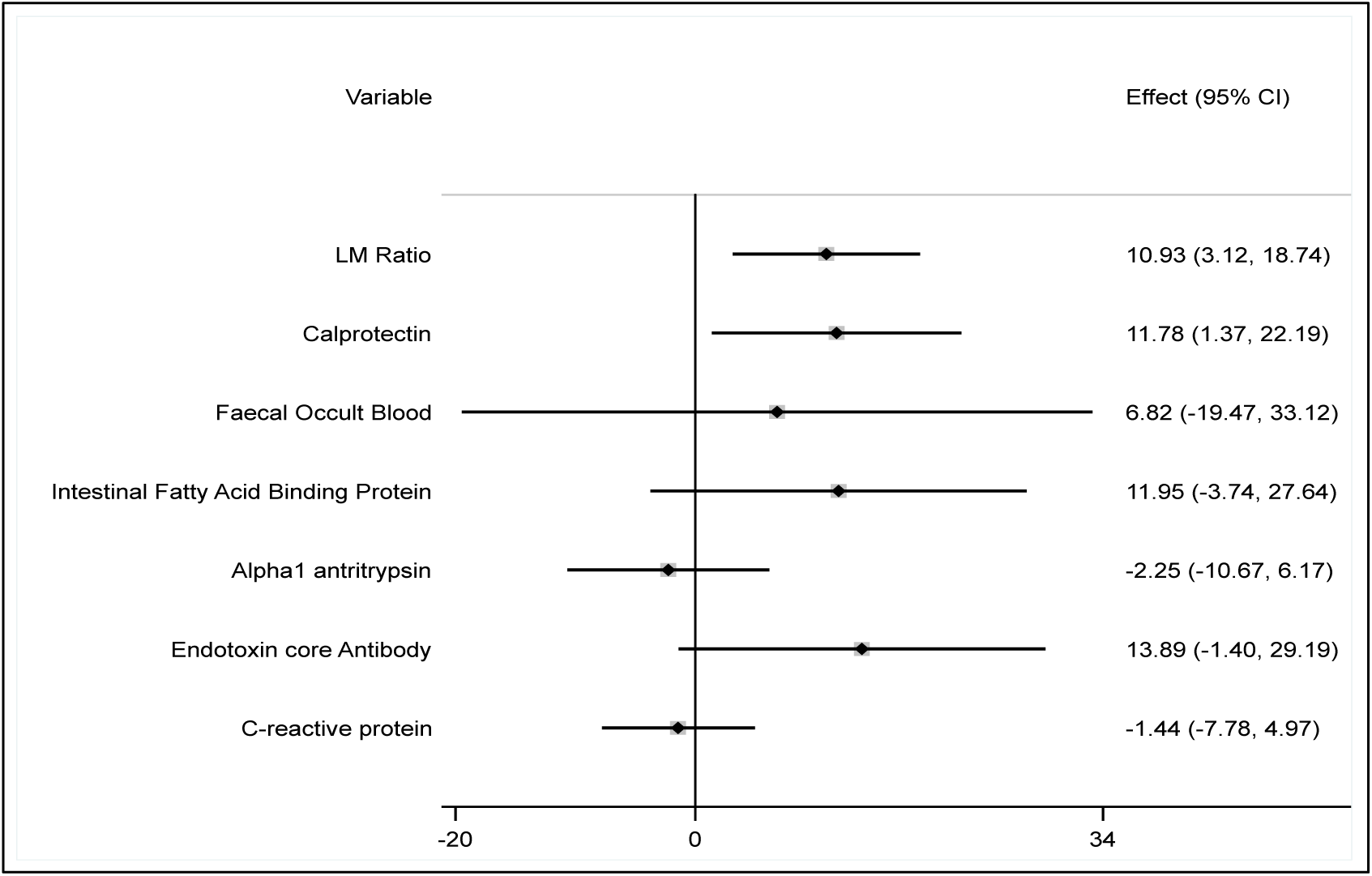
Forest plot showing association of time to maximum concentration (T_max_) of praziquantel with EED biomarkers after adjusting for age, sex and dose of PZQ. For each variable in the column on the left, the β regression coefficients are represented by the horizontal lines with the box being the effect size and the line on either side the 95% confidence intervals (CI). This is further presented as numerical values in the column on the right. Variables whose 95% CI do not cross the zero line on the x-axis are statistically significant.

### Enantiomer-specific analysis

S-PZQ exposure consistently exceeded that of R-PZQ across all metrics, with a median R/S AUC_inf_ ratio of 0.25, showing that the pharmacologically active R-enantiomer accounted for roughly a quarter of total PZQ exposure (***Table S1***). The R/S AUC ratio was higher in the 80 mg/kg group than in the 40 mg/kg group (median 0.34 vs 0.23, ***Table S2***), possibly due to partial saturation of stereoselective first-pass extraction at the higher dose. CRP was associated with higher exposure of both R-PZQ and S-PZQ to a near-identical degree (AUC β = 0.16 and 0.13, both p < 0.001) and showed no association with the R/S ratio (p = 0.22). In contrast, calprotectin and AAT were positively associated with the R/S AUC ratio (β = 0.08, p = 0.017 and β = 0.06, p = 0.008, respectively), indicating that greater intestinal inflammation and intestinal permeability was accompanied by proportionally higher exposure to the active R-enantiomer (***Table S3***).

### Absorption kinetics

Of the 91 participants in the 40 mg/kg group, 86 had reliable estimates, of which 44 (51%) displayed flip-flop kinetics (k_a_ < k_e_), implying absorption was slower than elimination. Absorption was correspondingly slower in flip-flop participants (median k_a_ 0.18 h⁻¹, absorption half-life = 3.9 h) than in those with conventional kinetics (k_a_ 0.45 h⁻¹, 1.6 h; ***Table S4***). None of the biomarkers reached statistical significance, but calprotectin showed a consistent trend across analyses: flip-flop participants tended to have higher median calprotectin than those with conventional kinetics (143 vs 94 µg/g; Mann-Whitney p = 0.096; ***Table S5***), and each log-unit increase in calprotectin trended toward higher odds of flip-flop kinetics (OR 1.43, 95% CI 0.95-2.14, p = 0.088; OR 1.45 after adjustment for age and sex). In the continuous analysis (linear regression of log k_a_), higher calprotectin similarly trended toward a lower absorption rate constant (β = −0.13, p = 0.074), a trend that disappeared in the k_a_ > k_e_ subset (β = 0.007, p = 0.82). No other biomarker showed even a trend toward an association with flip-flop status.

## Discussion

This study investigated the association between PZQ PK outcomes, namely, AUC, C_max_ and T_max_ and markers of EED in preschool age children treated for *S. mansoni* infection. Biomarkers representing intestinal pathology (FOB, plasma I-FABP, urine LM ratio, stool calprotectin, stool AAT), as well as microbial translation and systemic inflammation (EndoCAb and CRP) were assessed. Our findings indicate that higher CRP was independently associated with higher AUC and C_max_ while increased T_max_ was associated with higher LM ratio and calprotectin. Higher R-PZQ/S-PZQ AUC ratio was also associated with elevated calprotectin and AAT.

Faecal calprotectin is a cytosolic protein mainly from neutrophils [30] and is considered an intestinal inflammatory marker in various conditions such as inflammatory bowel disease [31] and schistosomiasis [32]. A higher urine LM ratio indicates both increased intestinal permeability (lactulose in urine) and decreased absorption (mannitol in urine), which results from disruption of the epithelium from inflammation-induced injury [33]. Diarrhoeal diseases also affect drug absorption as a result of pathological changes in the intestinal epithelium [34]. Two scenarios may explain higher T_max_ associated with elevated calprotectin and LM ratio. First, intestinal inflammation leads to destruction of absorptive enterocytes, which may explain the manifestation of malabsorption syndrome in EED [19]. Second, inflammation is also associated with downregulation of proteins involved in transport of molecules such as drugs through the intestinal epithelium [35], though the existence of such transporters for PZQ is yet to be described. Both scenarios point to slower PZQ absorption with greater intestinal inflammation.

Roughly half of the 40 mg/kg group (44 of 86) showed flip-flop kinetics, in which the terminal phase of the concentration-time profile reflects ongoing absorption rather than elimination, complicating the interpretation of elimination half-life. Of the biomarkers tested, only calprotectin showed a consistent trend toward this transition, although it did not reach significance (OR 1.43, p = 0.088). The signal was specific to the switch into flip-flop kinetics rather than graded slowing of absorption, as the association with k_a_ was lost when flip-flop participants were excluded (β = 0.007, p = 0.82). This pattern is mechanistically plausible: inflammation-associated slowing of gastrointestinal transit would delay delivery of PZQ to the absorptive epithelium, lowering the apparent absorption rate below the elimination rate. However, given the sparse sampling and the absence of significant associations, these observations should be regarded as hypothesis-generating.

Both the prolonged T_max_ and the high prevalence of flip-flop kinetics indicate slower absorption, which would be expected to reduce C_max_ and AUC. Elevated CRP, by contrast, was associated with higher C_max_ and AUC. CRP is an acute phase protein produced by the liver in response to immunogenic trigger broadly, [36, 37] but can also be produced by mesenteric adipocytes during intestinal inflammatory conditions [38]. Microbial translocation, a hallmark of EED, results in systemic inflammation and elevated CRP levels [19]. The CYP enzymes CYP1A2, CYP3A4 and CYP2C19 that contribute to PZQ metabolism are downregulated during inflammation as determined by serum CRP levels [20, 21]. The subsequent slower metabolism of PZQ during inflammation leads to higher plasma concentrations for a longer time, resulting in higher C_max_ and AUC. Reduced metabolism also leads to accumulation of a given drug and may be associated with increased toxicity [35]. High AUC of PZQ from population PK model of children in Uganda was associated with higher cure rates [14]. Decreased clearance of voriconazole has also been reported in association with elevated CRP, [39] in line with our findings.

Analysing the enantiomers separately localised these effects. CRP was associated with higher exposure of both R-PZQ and S-PZQ to a similar degree and was not associated with the R/S ratio. R-PZQ is metabolised primarily by CYP1A2 and S-PZQ primarily by CYP3A4, [3, 14] so isoform-selective suppression of hepatic metabolism would shift the ratio. The absence of a shift indicates that systemic inflammation raises both enantiomers in parallel, likely through a generalised reduction in first-pass metabolism. In contrast, calprotectin and AAT were associated with a higher R/S ratio. This is consistent with a gut-level effect, such as reduced intestinal CYP3A activity or altered epithelial transport, that preferentially increases exposure of the active R-enantiomer. These observations indicate that systemic and intestinal inflammation therefore act on different components of PZQ PK: the former on overall exposure, the latter on the relative exposure of the two enantiomers. Overall, inflammation also has an inverse relationship with the protein binding capacity, [40] implying high concentration of free drug in circulation. This could further explain the association of higher C_max_ and AUC with elevated CRP. Moreover, the high plasma concentration of free drug because of poor protein binding, as may occur during inflammation [35] could potentially lead to likelihood of toxicity. This observation is significant in the context of PZQ which is highly protein bound [3].

Contrary to the potential positive effects of increased C_max_ and AUC on efficacy due to increased drug exposure, increased T_max_ has a potential to reduce efficacy of a given drug because reduced rate of absorption affects the steady state concentration, [16] and may lead to exposure to subtherapeutic concentrations. This may not only affect cure rates but could also induce drug resistance.

This study did not control for other factors such as genetic polymorphisms of CYP enzymes. We did not assess treatment outcomes, so whether the observed changes in PZQ PK translate into altered cure rates remains to be determined. Finally, the four sampling time points limit the precision of the derived absorption parameters. The absorption rate constant and the classification of flip-flop kinetics should therefore be interpreted as exploratory.

In conclusion, intestinal inflammatory markers attributed to EED are associated with reduced rate of absorption of PZQ while systemic inflammation also has the potential to increase exposure to the drug. These findings represent the first report linking PK outcomes for PZQ to inflammation and add another dimension of potential factors that may explain the high variability in PZQ treatment outcomes. This study may also have implications for other oral therapeutics given in settings where EED is prevalent.

## Data availability

The anonymized data from the study are available as open data via LSHTM Data Compass, the London School of Hygiene and Tropical Medicine online data repository: https://doi.org/10.17037/DATA.00005306

